# Are we allowed to visit now? Concerns and issues surrounding vaccination and infection risks in UK care homes during COVID-19

**DOI:** 10.1101/2021.05.20.21257545

**Authors:** Clarissa Giebel, Kerry Hanna, Jacqueline Cannon, Paul Marlow, Hilary Tetlow, Stephen Mason, Justine Shenton, Manoj Rajagopal, Mark Gabbay

**Affiliations:** Department of Primary Care & Mental Health, University of Liverpool, Liverpool, UK; NIHR ARC NWC, Liverpool, UK; Lewy Body Society, Wigan, UK; Palliative Care Unit, University of Liverpool, Liverpool, UK; Sefton Advocacy, Sefton, Liverpool; Lancashire & South Cumbria NHS Trust

**Keywords:** Dementia, COVID-19, vaccination, social care, care homes, staff

## Abstract

**Background:** Vaccination uptake in the UK and increased care home testing are likely affecting care home visitation. With scant scientific evidence to date, the aim of this longitudinal qualitative study was to explore the impact of both (vaccination and testing) on the conduct and experiences of care home visits.

**Methods:** Family carers of care home residents with dementia and care home staff from across the UK took part in baseline (October/November 2020) and follow-up interviews (March 2021). Public advisers were involved in all elements of the research. Data were analysed using thematic analysis.

**Results:** Across 62 baseline and follow-up interviews with family carers (n=26; 11) and care home staff (n=16; 9), five core themes were developed: Delayed and inconsistent offers of face-to-face visits; Procedures and facilitation of visits; Frustration and anger among family carers; Variable uptake of the COVID-19 vaccine; Misinformation, education, and free choice. The variable uptake in staff, compared to family carers, was a key factor seemingly influencing visitation, with a lack of clear guidance leading care homes to implement infection control measures and visitation rights differently.

**Conclusions:** We make five recommendations in this paper to enable improved care home visitation in the ongoing, and in future, pandemics. Visits need to be enabled and any changes to visiting rights must be used as a last resort, reviewed regularly in consultation with residents and carers and restored as soon as possible as a top priority, whilst more education needs to be provided surrounding vaccination for care home staff.

## Background

Care homes have been affected to the greatest extent by the COVID-19 pandemic, heightened by the fact that residents are most susceptible to the virus. In 2021 alone, over 10,000 care home residents in England have passed away from COVID-19 (ONS database, 2021).

There is emerging quantitative evidence on COVID-19 outbreaks in care homes and management of infection risks (Burton et al., 2020; 2021), with a body of research into the effects on health care staff yet limited evidence on the impact on social care staff (De Kock et al., 2021; Hanna, Rapa, Dalton et al., 2021). In a recent international report by Low-Fay (2021), summarising the limited available evidence into the effects of the pandemic on residents, family carers, and staff, the authors made strong recommendations for safe visiting to be enabled immediately, to ensure improved well-being for all involved. Social engagement is vital (Sommerlad et al., 2019), and research into lack of social engagement in the community for people with dementia has already shown detrimental impacts on faster deterioration (Giebel et al., 2020). The negative impact of lack of social engagement has also been shown in the care home setting (Ayalon & Avidor, 2020; van der Roest et al., 2020).

Guidance surrounding care homes and visitation, and infection control, where they are available (Giebel et al., under review), are changing rapidly. This is particularly the case more recently due to the large vaccination rollout across the UK, with over half of the population vaccinated with a first dose in April 2021. Care home residents and health and social care staff were prioritised in accessing the vaccine, as well as family carers, with reports of reduced vaccination rates among social care staff. A survey into vaccine hesitancy in Liverpool-based care home staff showed that on average only half of care home staff in each care home (51.4%) had been vaccinated, with concerns about lack of vaccine research, misinformation about fertility side effects, and being off-site stated as common reasons for vaccine hesitancy (Tulloch et al., preprint). However, data are based on a survey with only 50% of approached Liverpool-based care homes responding, and are specific to this region within North West England, with no qualitative data to date on vaccination of care home staff across the UK or other countries. This is important to understand however, as social care staff provide care to some of the most vulnerable members of our society, in particular those groups who are most vulnerable to infection and mortality from COVID-19 (Daras et al., 2021). Moreover, there is no research to date showcasing the impact of vaccination rollout and increased testing across care homes and family carers on visitation, whilst we know that testing in care homes proves to be an effective measure of infection control (Micocci et al., 2021).

In March 2021, the government made two announcements and allowed at first one essential visitor into the care home (8^th^ of March), which was followed by a second essential visitor (29^th^ of March). Different countries have different regulations, and in the Netherlands for example there is a law against blanket care home closures to family and friends. This is also currently being debated in the UK.

The aim of this study was to explore the longitudinal impact of the pandemic on care home visiting rights and the effects of vaccination and testing on visitation. With no evidence to date, this study will provide crucial first findings with potential implications for care home visitation guidance.

## Methods

### Participants and Recruitment

Family carers who have a relative with dementia residing in a care home, and care home staff, were eligible to take part. Participants were included if they were aged 18+ and were residing/working in the UK Initial recruitment took place via advertisement on social media and third sector organisations in October 2020. Participants were purposefully sampled for follow-up interviews in March 2021, in order to gather longer-term experiences of both family carers and the care home workforce, following significant public health changes at that time such as, the COVID-19 vaccination rollout in the UK and a further national lockdown.

Ethical approval was obtained from the University of Liverpool ethics committee (Ref: 7626) prior to study commencement, and an amendment later granted for the follow-up interviews.

### Data and data collection

Baseline interviews were conducted between October and November 2020, and follow-up interviews were conducted in March 2021, when vaccinations were ongoing and restrictions were lifted to allow first one, then two visitors into the care home and hold hands using PPE. Figure 1 shows a timeline of care home restrictions in comparison with national changes, whereby it is important to highlight that each nation (England, Wales, Scotland, Northern Ireland) imposed their own restrictions from summer 2020.

**Figure 1.**
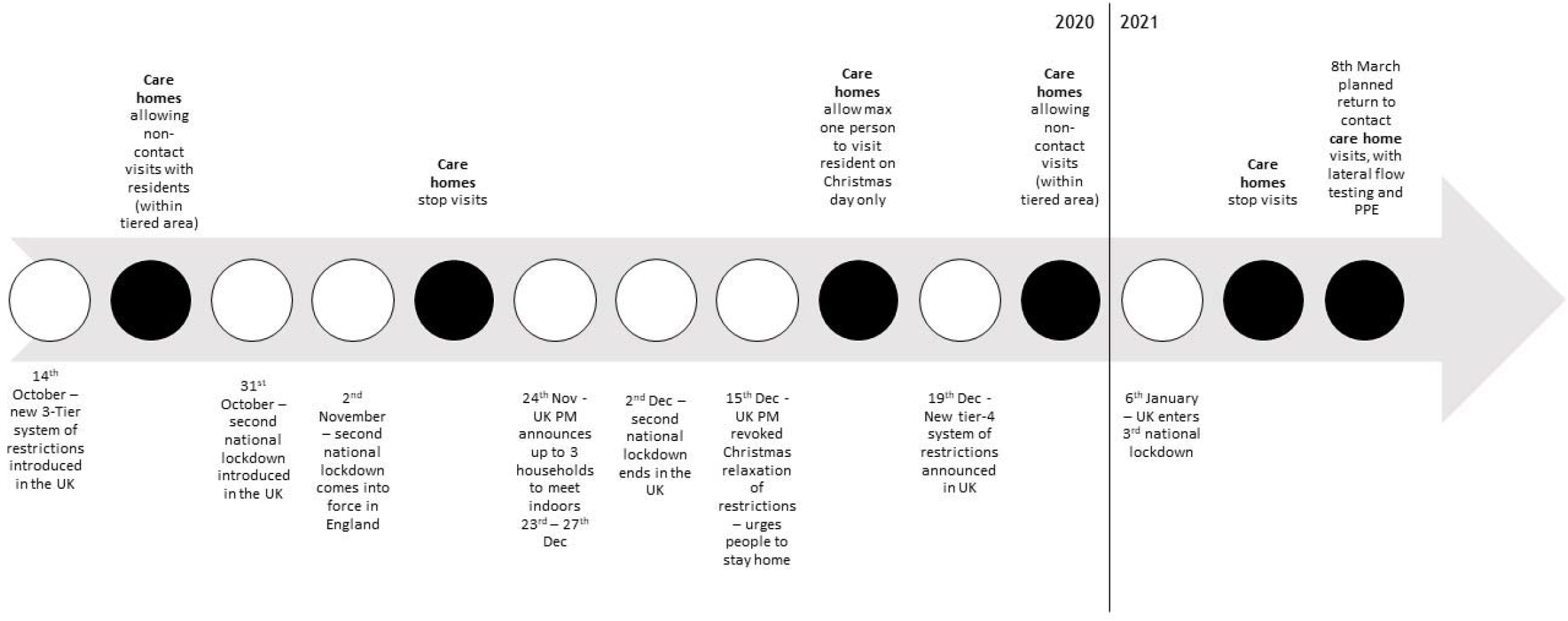
Timeline of UK public health restrictions in the time of COVID-19, from October 2020 – March 2021. White circles indicate UK restrictions, and black circles indicate care home restrictions in response to the public health measures at that time.

We collected basic demographic characteristics of participants including age, gender, ethnicity, as well as relationship with relative with dementia and dementia subtype from family members, and years of working in the care home sector, staff role, and size of the care homes from care home staff.

The interview topic guides for baseline and follow-up interviews were co-produced with clinicians, unpaid carers of people living with dementia and academics. Iterations of the topic guide were circulated between team members until a final version was agreed. During baseline interviews, participants were asked about changes to their caring roles since the pandemic, their experiences of viral testing and COVID-19 safety measures employed in the care home, resident visits and communications between family members the care home, and the impact of the restrictions on the staff and residents. Follow-up interview questions centred on changes to participants’ experiences and restriction impacts over time, and included further questions about their experiences of and views about COVID-19 vaccination and vaccine uptake in care home residents and staff and for themselves as carers, as well as changes to visiting arrangements.

Semi-structured, remote interviews were conducted, with participants offered their preferred form of communication (phone or online). Interviews were audio-recorded, with verbal consent obtained and recorded at the beginning of each interview. Audio files were transcribed and in the process anonymised. The average length of interview at baseline was 29 (+/- 11) minutes, [12-58], and at follow-up was 24 (+/-7) minutes [13-37].

### Data analysis

Transcripts were coded using thematic analysis (Braun & Clarke, 2006). Baseline interviews were coded shortly after data collection, and follow-up interviews were coded separately shortly after follow-up data collection. Data saturation was observed in the baseline interviews, and interviews ceased after interview 42. In the follow-up interviews, data saturation was suspected after interview 18, but the following two interviews were honoured as these had already been arranged with the participants, and saturation was confirmed.

Using thematic analysis, five research team members all experienced in qualitative analysis, including one former carer trained up in data analysis (JC), coded the transcripts. Specifically, each transcript was coded individually by two research team members before meeting and discussing developing themes and codes. In the first meeting, we had coded three quarters of transcripts and discussed the findings to help with subsequent conceptualisation of themes. After the final analysis meeting, final themes were presented to all team members, highlighting both an inductive and subsequent deductive analysis approach.

### Public involvement

Three carers (two former, one current) were active team members and involved in all aspects of the study, from conceptualisation of the project, to designing study documents, helping interpreting findings, and dissemination. One carer was also involved in the analysis of the data. Public advisers were reimbursed according to NIHR INVOLVE guidelines.

## Results

We conducted a total of 62 interviews (42 baseline and 20 follow-up interviews). At baseline, 26 family carers and 16 care home staff participated. At follow-up, 11 family carers and 9 care home staff participated. Across all baseline interviewees, the majority were female (n=31), White British (n=35) and with a mean age of 54.8 (±15.9). The majority of participants resided in the least disadvantaged quintile (IMD=1) as reported from their postcode IMD score. Of the 26 family carers recruited, the majority were adult children (n=16), with the remaining relations spouse or partner. The most common dementia subtype, of the PLWD residing in a care home, was Alzheimer’s (n=8), followed by Lewy Body (n=6) and Vascular (n=4). Of the 16 care home staff, the mean years of working in a care home was 9.3 (±10.6), with care assistant and manager the most common job roles (n=4 respectively). Table 1 shows the full demographics of the recruited participants in both the baseline and follow-up interviews.

**Table 1.** Demographic characteristics of family carers and care home staff.

|  | Family carers<br>baseline (n=26)<br>N (%) | Family carers<br>follow up (n=11)<br>N (%) | Care home staff<br>baseline (n=16)<br>N (%) | Care home staff<br>follow up (n=9)<br>N (%) | Total sample<br>(n=42)<br>N (%) |
| --- | --- | --- | --- | --- | --- |
| Gender |  |  |  |  |  |
| Female | 18 (69.2%) | 8 (72.7%) | 13 (81.3%) | 8 (88.9%) | 31 (73.8%) |
| Male | 8 (30.8%) | 3 (27.3%) | 3 (18.8%) | 1 (11.1%) | 11 (26.3%) |
| Ethnicity |  |  |  |  |  |
| White British | 22 (84.6%) | 10 (90.9%) | 13 (81.3%) | 7 (77.8%) | 35 (56.5%) |
| White Other | 2 (7.7%) | 1 (9.1%) | 1 (6.3%) | 1 (11.1%) | 3 (4.8%) |
| BAME | 2 (7.7%) | 0 | 1 (6.3%) | 0 | 3 (4.8%) |
| Prefer not to say | 0 | 0 | 1 (6.3%) | 1 (11.1%) | 1 (1.6%) |
| Relationship with PLWD |  |  |  |  |  |
| Spouse | 9 (34.6%) | 3 (27.3%) |  |  |  |
| Partner | 1 (3.8%) | 0 |  |  |  |
| Adult child | 16 (61.5%) | 8 (72.7%) |  |  |  |
| Dementia subtype |  |  |  |  |  |
| Alzheimer's disease | 8 (30.8%) | 4 (36.4%) |  |  |  |
| Mixed dementia | 2 (7.7%) | 0 |  |  |  |
| Vascular dementia | 4 (15.4%) | 2 (18.2%) |  |  |  |
| Lewy Body dementia | 6 (23.1%) | 4 (36.4%) |  |  |  |
| Other | 2 (7.7%) | 1 (9.1%) |  |  |  |
| Unknown | 4 (15.4%) | 0 |  |  |  |
| IMD Quintile<sup>2</sup> |  |  |  |  |  |
| 1 (least disadvantaged) | 11 (42.3%) | 6 (66.7%) | 3 (23.1%) | 2 (28.6%) | 14 (43.8%) |
| 2 | 4 (14.5%) | 2 (22.2%) | 3 (23.1%) | 2 (28.6%) | 2 (21.9%) |
| 3 | 0 | 0 | 3 (23.1%) | 2 (28.6%) | 3 (9.4%) |
| 4 | 3 (11.5%) | 0 | 1 (7.7%) | 0 | 4 (12.5%) |
| 5 (most disadvantaged) | 1 (3.8%) | 1 (11.1%) | 3 (23.1%) | 1 (14.3%) | 4 (12.5%) |
| Job role |  |  |  |  |  |
| Activity Coordinator |  |  | 1 (6.3%) | 0 |  |
| Care home liaison |  |  | 1 (6.3%) | 1 (11.1%) |  |
| Care quality |  |  | 1 (6.3%) | 0 |  |
| Care assistant |  |  | 4 (25.0%) | 3 (33.3%) |  |

**NOTE: This preprint reports new research which has not yet been peer reviewed.**

|  |  |  |  |  |  |
| --- | --- | --- | --- | --- | --- |
| Senior care assistant |  |  | 2 (12.5) | 0 |  |
| Night care assistant |  |  | 1 (6.3%) | 1 (11.1%) |  |
| Housekeeper |  |  | 1 (6.3%) | 1 (11.1%) |  |
| Matron |  |  | 1 (6.3%) | 0 |  |
| Manager |  |  | 4 (25.0%) | 3 (33.3%) |  |
|  | **Mean (SD), [Range]** |  |  |  |  |
| Age<sup>1</sup> | 62.3 (±9.5) [42-89] | 61.1 (±5.2) [51-68] | 41.8 (±16.6) [18-62] | 43.3 (±17.2) [21-60] | 54.8 (±15.9) [18-89] |
| Years of education | 17.9 (±2.9) [11-23] | 18.09 (±1.5) [16-20] | 15.7 (±2.7) [11-20] | 16.4 (±2.6) [11-19] | 17.1 (±3.0) [11-23] |
| Care home capacity | 41.5 (±17.4) [18-76] | 38.9 (±18.2) [18-76] | 42.2 (±15.8) [12-64] | 49.7 (±11.6) [36-64] | 41.7 (±16.6) [12-76] |
| Years working in a care home |  |  | 9.3 (±10.6) [1-35] | 7.0 (±11.1) [1-35] |  |
| Years since dementia diagnosis | 6.7 (±3.6) [2-16] | 7.0 (±4.4) [2-16] |  |  |  |
| Years (PLWD) residing in a care home | 2.7 (±2.1) [1-10] | 2.8 (±1.9) [1-7] |  |  |  |

<sup>1</sup>n=1 care home staff = prefer not to say, <sup>2</sup>n=4missing data

### Qualitative findings

Thematic analysis identified five themes: (1) Delayed and inconsistent offers of face-to-face visits; (2) Procedures and facilitation of visits; (3) Frustration and anger in family carers; (4) Variable uptake of the COVID-19 vaccine; (5) Misinformation, education, and free choice. Table 2 includes key representative quotes from interviews by theme.

**Table 2.**
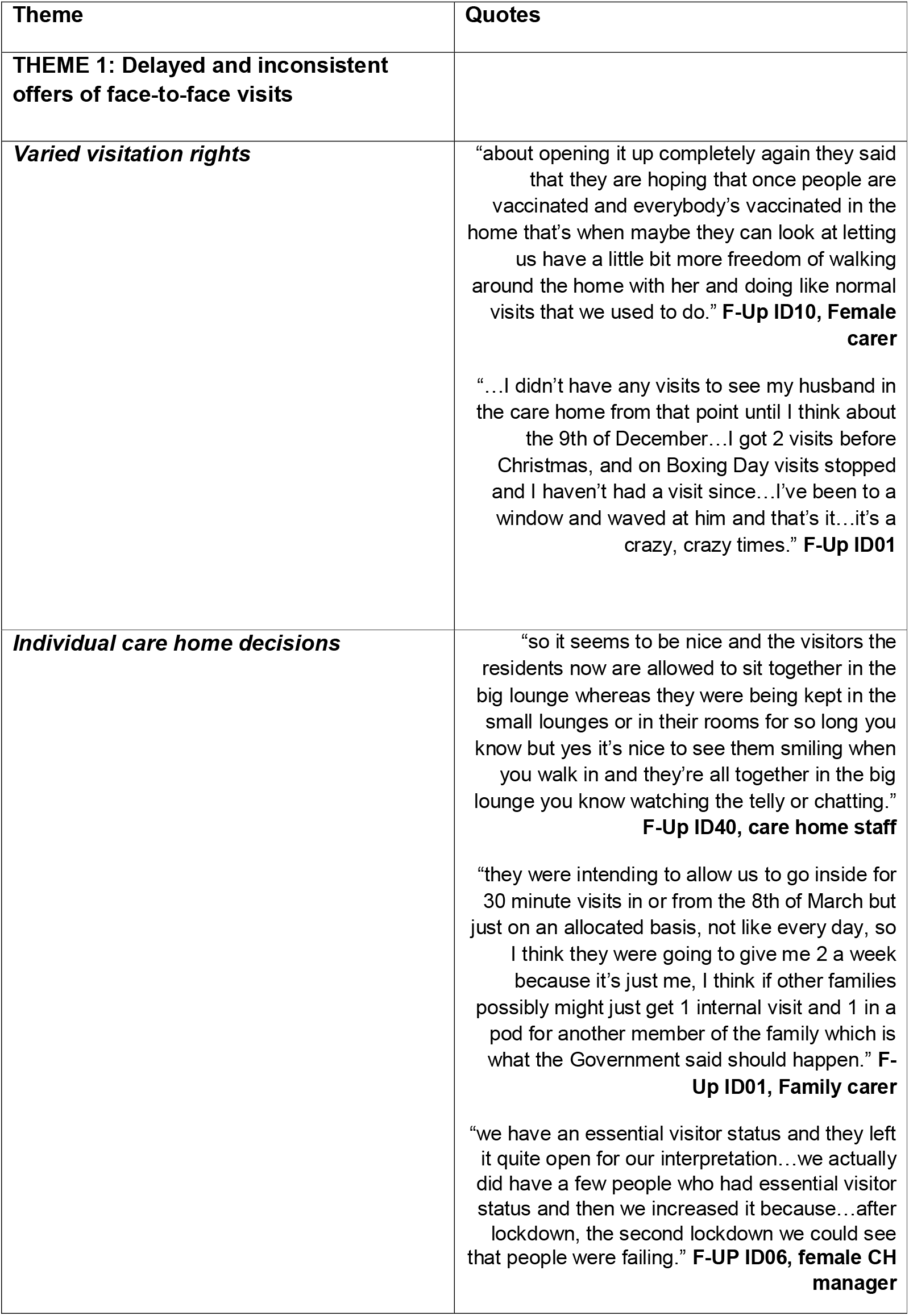
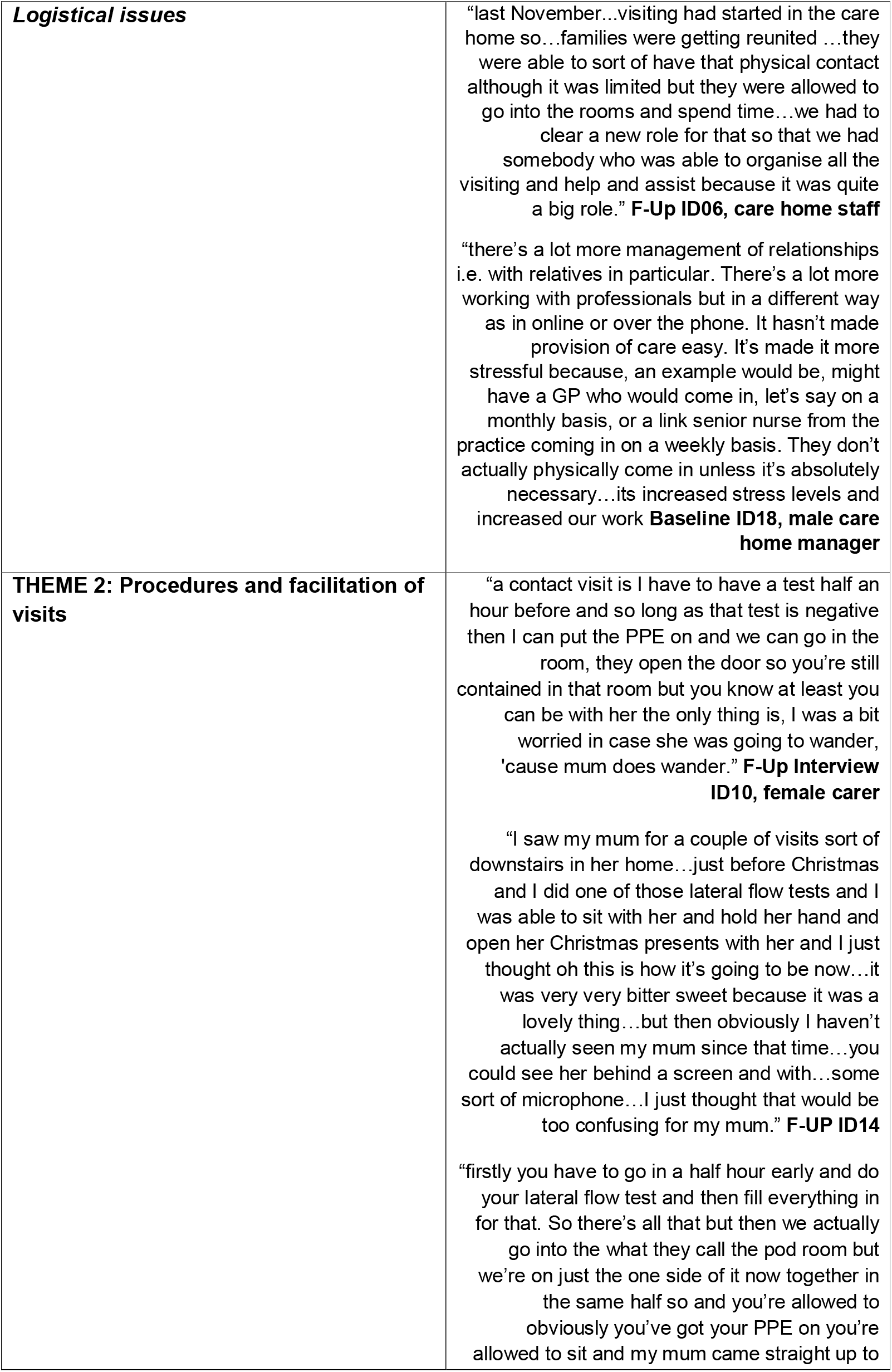
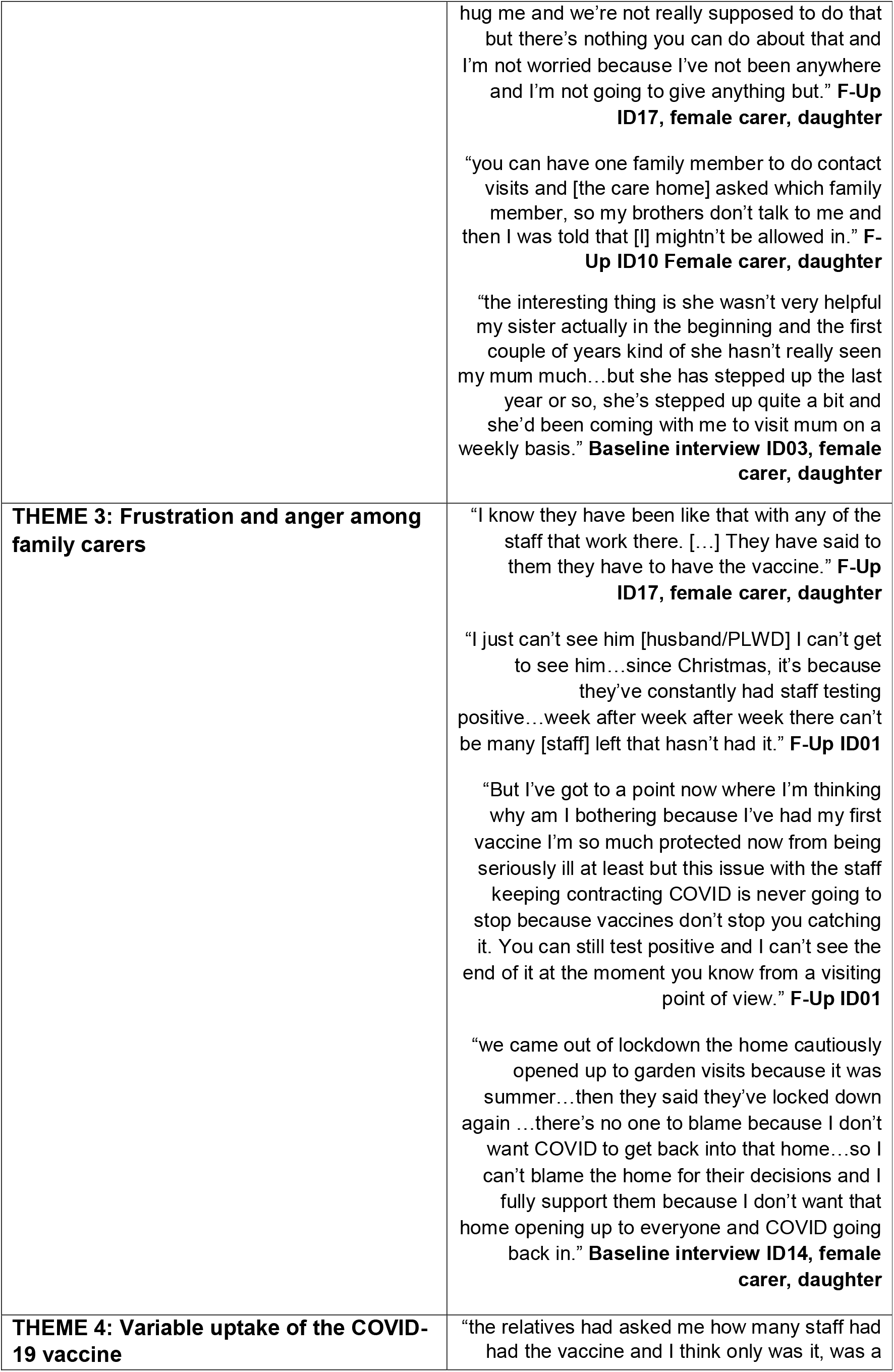
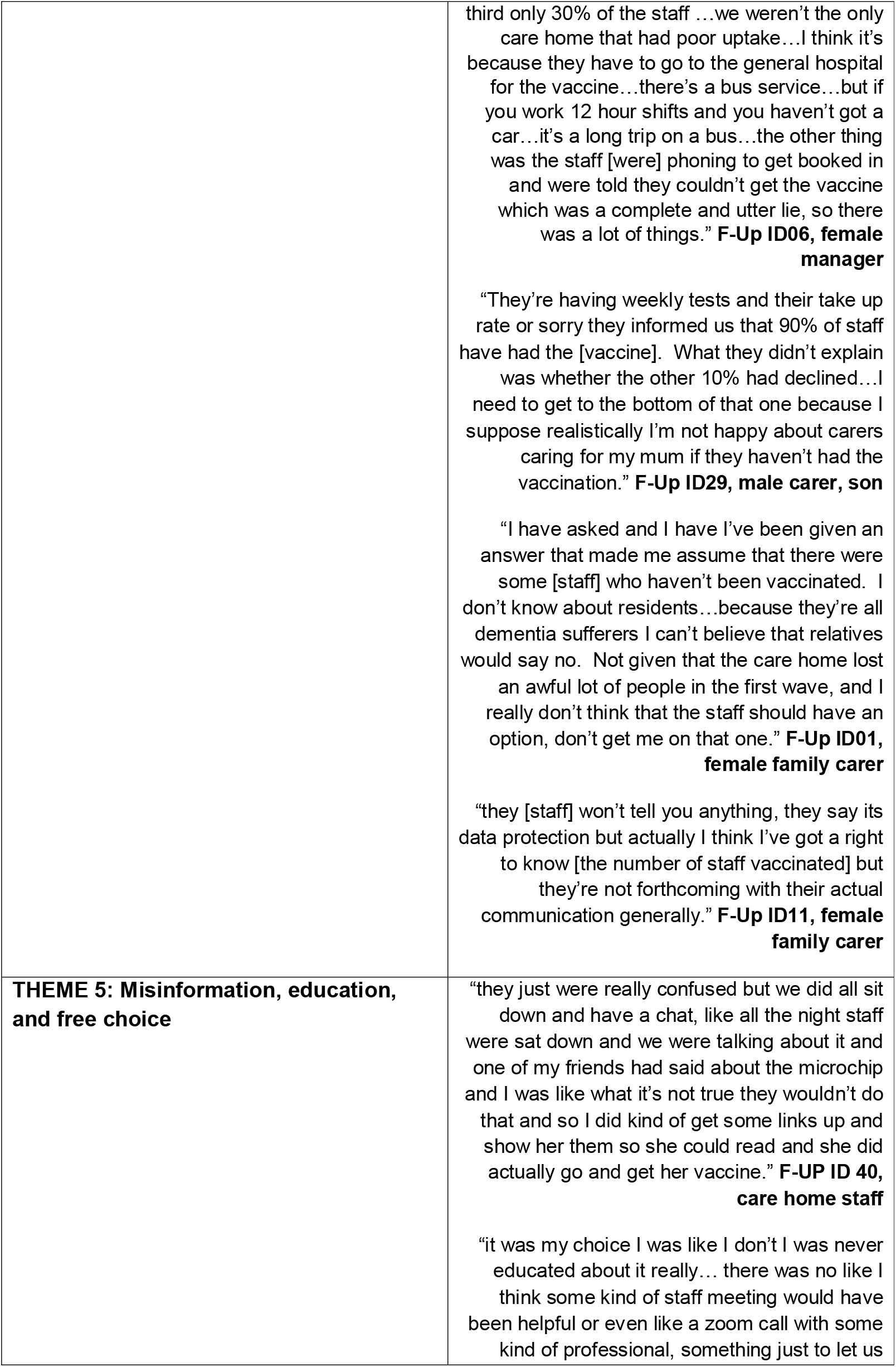
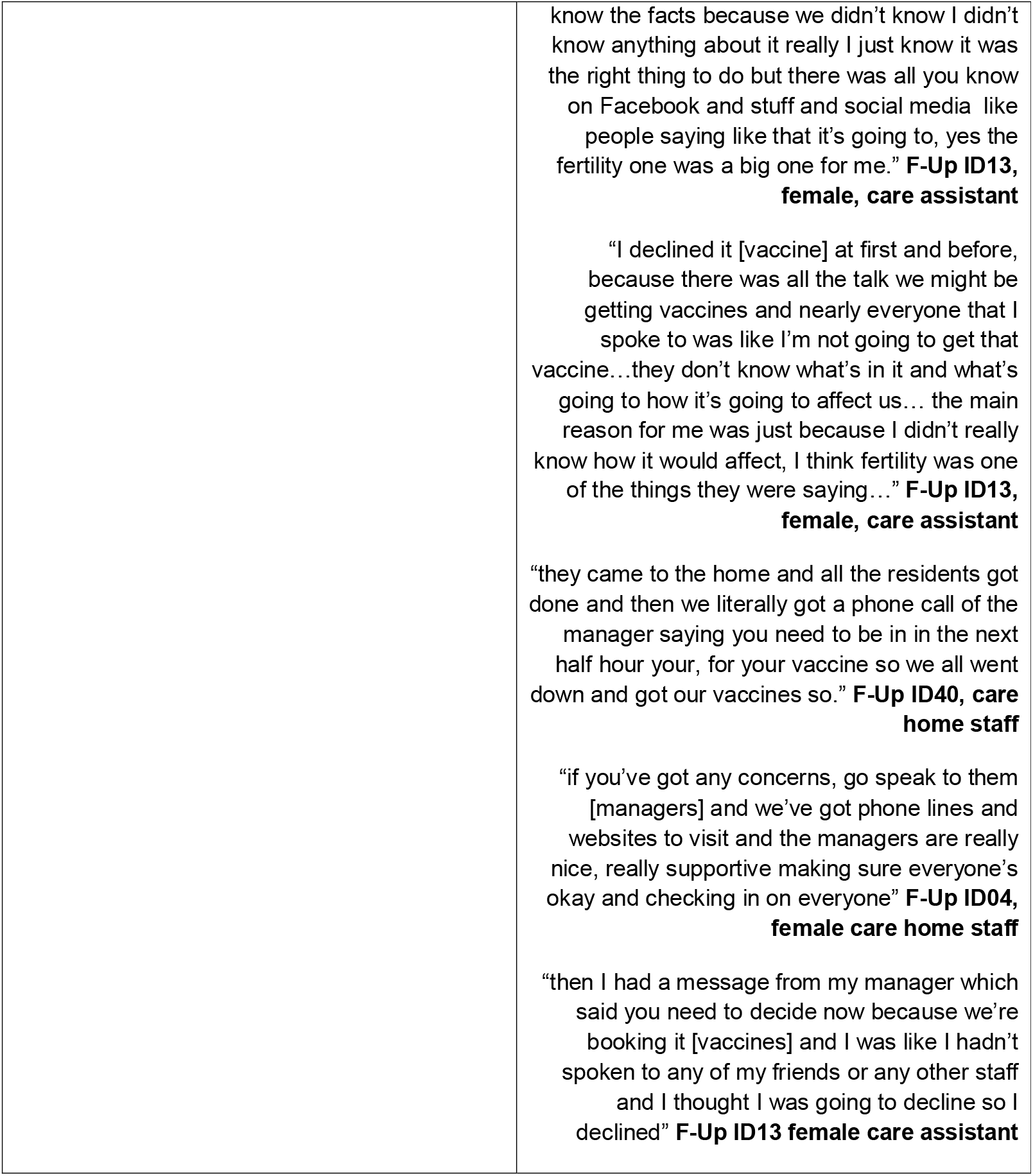
Interview quotes by theme.

### THEME 1: Delayed and inconsistent offers of face-to-face visits

#### Varied visitation rights

Family carers experienced varied visitation rights, with some carers only visiting briefly before Christmas and a general uncertainty as to when the care homes would open up for resuming normal face-to-face visits again. Some care homes appeared to be wary of enabling visits waiting for vaccinations to take place protecting everyone as much as possible prior to enabling visits again.

#### Individual care home decisions

Each care home appeared to implement the guidance in the way they saw best, with some homes enabling visits and to a greater extent, whilst others were more reluctant to enable visits. This was reflected in some staff not wanting family carers coming into the home for fear of virus transmission, albeit some staff themselves had not been vaccinated, thereby increasing the risk of infection transmission. Family carers were thus unable to control visitation, regardless of being vaccinated themselves, and were reliant on each care home to make its own decisions in terms of visitation.

#### Logistical issues

Increased testing and other infection control measures, arranging visits, and attending alternative face-to-face visits all add additional workload for care home staff. Staff highlighted the additional demands this places on their time, which results in less ward time to care for residents. In some care homes, additional staff had been recruited specifically for these COVID-19 related measures, whilst in others staff had to be taken off the ward and could not care for the residents. This could lead to some care homes being better placed at enabling more visits, whilst others did not have the staff capacity to enable many visits.

### THEME 2: Procedures and facilitation of visits

Family carers had mixed experiences surrounding visitation, with alternatives to face-to-face visits not beneficial for all due to dementia- and ageing-related issues of hearing, vision, and lack of understanding. The majority of visits which occurred were alternative face-to-face visits (pod and window visits). Care homes implemented strict testing, PPE, and distancing procedures, and family carers shared the lengthy logistics of visiting their relatives, involving testing ahead of the visit and having to wait outside until the test comes back negative. Despite adhering to social distancing measures at the time of data collection and before (which changed during the period of data collection), one family carer recounted how her mother with dementia came up to her and hugged her, with the carer appreciating it was against the infection control measures but knowing she had done everything possible to minimise any infection risks.

The announcement of essential visitor status caused some difficulties for family carers where more than one person wanted to visit the relative at the home face to face. Some carers who were interviewed were not allocated the essential visitor status but their sibling or other relative instead. This could cause issues within the family, particularly when relatives did not speak with one another and were not receiving any information from the one family member who was allowed to visit face-to-face under new guidance. Compared to pre-pandemic and earlier pandemic experiences, which showed some family support growing amongst relatives, the new announcements caused potential rifts amongst relatives in deciding who was allowed to visit.

### THEME 3: Frustration and anger among family carers

Family carers were angry and frustrated about the fact that they had not been able to enter care homes and have face-to-face visits with their relative despite being tested, vaccinated and careful about adhering to restrictions. This was aggravated by family carers seeing care home staff sitting next to their relative behind a pod screen or window, in close proximity, without much knowledge as to whether staff had been vaccinated or not. In addition, on each alternative visit, different members of staff could be facilitating the visit sitting close to the resident, thereby again increasing potential infection risk when family carers had often little to no knowledge communicated to them about vaccination of staff.

Some family carers were frustrated as staff tested positively sequentially over prolonged periods of time, leading to care home closures and family carers unable to visit their relatives, and to some extent blaming staff for not being able to enter the care home, whilst family carers themselves were in strict adherence of the regulations.

Family carers reported empathy and understanding in the earlier baseline interviews, in contrast to the above accounts depicting frustration and exacerbation in response to the unvaccinated care home workforce. This contrast of emotions appears to relate to the persistent COVID-19 outbreaks in homes, and subsequent visiting restrictions, despite the fact that family carers cannot enter the home in any form during the national lockdown. Therefore, the care home staff were viewed as solely responsible for virus transmission, and thus, the reason for persistent lockdown with homes prohibiting contact visits restarting.

### THEME 4: Variable uptake of the COVID-19 vaccine

There was a variable uptake in vaccination between family carers and care home staff. All family carers had been vaccinated, whilst not all staff had or colleagues within the same care home had not been vaccinated. Vaccination was overall also slower in staff, according to family carers, than amidst themselves, who had all been keen to get vaccinated.

Variations in, and reduced uptake of, vaccination in staff compared to family carers was cause for concern, with some family carers actively questioning why some staff had not been vaccinated. This was however only the case where family carers knew about the rate of staff vaccination at the homes, which often was not the case, showcasing a wider lack of communication between care homes and families.

### THEME 5: Misinformation, education, and free choice

Care home staff reported different issues surrounding information about COVID-19 vaccination, with some of their co-workers misinformed about the vaccine. There appeared to be a lack of credible information and sufficient education to fully inform staff about getting vaccinated, with side effects on fertility as well as being microchipped mentioned as reasons for not getting vaccinated. Misinformation seemed to have spread via social media and the internet more broadly. Interviewed staff reported, however, that they spoke with their colleagues about misinformation and talked through their concerns, which led some colleagues to get vaccinated in the end, as well as themselves where the participants were initially reluctant to.

Staff also reported logistical barriers getting vaccinated, with too little notice provided ahead of the date of vaccination, and no alternative offered. In light of some misinformation, these short windows of time seemed to hinder some staff further in accessing the vaccine, albeit the majority of interviewed staff claiming to be vaccinated.

Another key factor that influenced vaccination uptake amongst care home staff was the communication from their care home manager. Where the manager actively encouraged all staff to get vaccinated through education and open discussions, vaccination rates, according to interviewed staff, appeared to be higher than in homes where managers were not expressing an opinion.

## Discussion

This is the first study to have explored the impact of vaccination and increased testing on care home visitation. Findings showed how visits were often delayed and inconsistent, and subject to various barriers and implementation of guidelines dependant on each care home. The variability in vaccination between staff and family carers appeared to be the biggest barrier of all, and cause of considerable angst for family carers.

There has been no consistent approach to enabling care home visits during this pandemic. Our findings show that due to a lack of clear guidance, each care home interpreted the guidance themselves and made decisions on how and when visits could be enabled. This left some family carers seeing their relatives via alternative face-to-face visits, whilst others were experiencing delays. This was particularly the case with the March announcement of essential visitors being allowed into English care homes, with care homes not having received any communication from the government about these changes in advance. Social contact and enabling family visits are vital to relatives and residents however, and a human right (Butchard & Kindermann, 2019). As Lee-Fay and colleagues (2021) reported in an international overview of care home recommendations in the time of the pandemic, safe visiting needs to be enabled, which is corroborated by our findings whilst also highlighting the barriers which normally one would expect to act as facilitators.

Whilst family members were initially (at baseline) more understanding of the situation, albeit upset, the emotional intensity appeared to have changed by follow-up. Family carers were overall frustrated and angry at the strict measures in place for them in terms of visitation, compared to less strict measures on infection control for staff. Family carers were adhering to restrictions, and were all vaccinated and willing to test in order to see their relatives as soon as possible. Regardless, the vast majority were only allowed to see their relative behind a window or pod screen, whilst a different member of staff sat close to the relative on each of these alternative face-to-face visits. This was presumably heightened by the consistent lack of communication between care homes and families (Giebel et al., under review), increasing the emotional upset in family members. The pandemic is having a stark impact on people’s mental well-being already (Fancourt, Steptoe, Bu, 2021; Hanna et al., 2021), and the inability to visit loved ones can exacerbate low mental well-being in family members, as evidenced in emerging research (Ayalon & Avidor, 2020; van der Roest et al., 2020). Therefore, in addition to enabling visits, family members need better social and psychological support to cope with how the pandemic has affected their relationship with their relative, with peer support groups for example provide emotional and social benefits (Keyes et al., 2016). This could also be addressed by improved communication between the care home and families, allowing families to have a better insight into the well-being of their relative when visits can be difficult to access, and a more open discussion about the causes of restrictions and balance of risk.

Increased testing and availability of vaccination would be expected to be a facilitator of face-to-face visits. However, variability in vaccine uptake and logistics around planning visits can also act as barriers. There appeared to be misinformation among staff surrounding the consequences and side effects of vaccination. Fertility, microchips, and other apparent side effects were raised, which caused delays in some staff getting the vaccine. Whilst it is beneficial that most staff got vaccinated in the end, any delays in getting vaccinated can cause more time to get infected and to spread the virus, in a population that is one of the most vulnerable in our societies. In addition, this can delay the ability for family carers to visit as infection outbreaks may be more likely. Ladhani and colleagues (2020) for example reported increased infection risk in staff working across multiple care homes. The notion of vaccine hesitancy amongst social care staff is corroborated in a small survey in a small geographical region of the North West of England, reporting vaccine hesitancy in 50% of surveyed care home staff (Tulloch et al., 2021). Equally, lower education in the French working age population has been found to be related to lower vaccine uptake (Schwarzinger et al., 2021). Whist this showcases that vaccine hesitancy is not restricted to the social care taskforce, and there being a great deal of misinformation surrounding the pandemic in general (Green et al., 2021), UK social care staff overall has low educational entry requirements (House of Commons Committee of Public Accounts, 2018), which may be one of the reasons for vaccine hesitancy. However, there are many factors at play, and hesitancy, as indicated by Tulloch et al.’s (2021) findings, cannot easily be explained by one single reason. To overcome the issue of misinformation, more adequate education and information about the vaccines and the benefits need to provided, not just for this but also for future pandemics.

Logistical issues can also provide a barrier to vaccine uptake in care home staff, which in return can impact on decisions about infection control measures in care homes and allowing family members inside. We reported occasions where staff were informed less than an hour before the vaccinations were taking place, which leaves little time for staff to receive proper information about the vaccine and if still unsure, to ask sufficient questions. These difficulties in adequate vaccination rollout contribute to the anger experienced by family carers with fewer staff potentially vaccinated than if there was appropriate timing. This lack of advance notice seems to be supported by a general lack of guidance for care homes, as the announcements of essential visitor rights were equally not communicated to care homes in advance. This strongly highlights the general lack of communication and support from decision makers in managing infection control in the care home settings, so that care homes have to rely on their own judgements. Preliminary findings by Marshall et al. (2021) corroborate how care homes were left without much support and instead often supported themselves and received support from their communities.

Based on these novel findings, we make five specific recommendations for the care home sector, to the benefit of staff, family carers, and residents:

- Face-to-face visits are a human right and the right to see loved ones should only ever be removed as a last resort, regularly reviewed in consultation with residents and carers, and restored as a high priority as soon as possible
- Support for care homes for the effective implementation of infection control measures and access to personal protective equipment alongside health staff, to avoid taking staff time away from caring for residents and leading to stopping visits as a default, lower cost, protective response
- Need for better information support and guidance surrounding vaccination for all involved and improved logistical processes for vaccine delivery
- General need for better guidance and communication to support care home staff in their work delivery and communication between care homes and families
- Vaccination of social care staff should be mandatory

Whilst this study benefits from having captured the precise moment when visitation restrictions were officially eased for care homes in England, and being the first study to explore the impact of heightened infection control measures (testing, vaccination) on care home visitation, there were some limitations. This longitudinal study only interviewed family members of care home residents and care home staff, thereby only collecting some proxy information on how people with dementia residing in the care homes were faring. Considering pandemic restrictions of not collecting data in care homes, as well as the difficulty of obtaining experiential data from people who mostly lack capacity to consent, given their advanced dementia, this was the most feasible way of collecting data. Further research needs to explore impact of restrictions on residents’ well-being and functioning, which can be achieved via quantitative measurements. A positive of the sample is the fact that staff were recruited from 16 different care homes across the UK, thus broadening the representativeness of care home experiences. Additionally, our sample was lacking ethnic minority representation, and mostly included family carers and staff from a White ethnic background. In light of increased susceptibility of people from minority ethnic backgrounds to the virus (Daras et al., 2021), future research needs to explore their views, as ethnicity may affect behaviour and attitudes towards infection control measures and visitation.

## Conclusions

This study provides the first insights into how increased infection control measures (testing and vaccination) have impacted on care home visitation. The lack of social contact with relatives has been detrimental to family members, with our findings providing strong evidence-based recommendations for the continued handling of the COVID-19 pandemic going forward, as well as for other future infection outbreaks. With voices emerging on implementing a law to enable social contact with loved ones for residents, this study supports this notion, whilst more in-depth research is required on the precise impacts of residents from their points of views.

## Data Availability

Data may be made available after one year since data collection upon reasonable request.

## Acknowledgements

We wish to thank all family carers and care home staff who took part in this study, and we also wish to thank the many family carers who expressed an interest to take part after we were already booked up. We also wish to thank Maxine Martine and Lynne McClymont for transcribing the audio files very swiftly to analyse the data in time.

